# Performance of RT-PCR on saliva specimens compared to nasopharyngeal swabs for the detection of SARS-CoV-2 in children: A prospective comparative clinical trial

**DOI:** 10.1101/2021.02.27.21252571

**Authors:** Yves Fougère, Jean Marc Schwob, Alix Miauton, Francesca Hoegger, Onya Opota, Katia Jaton, Rene Brouillet, Gilbert Greub, Blaise Genton, Mario Gehri, Ilaria Taddeo, Valérie D’Acremont, Sandra A. Asner

## Abstract

**Background:** Saliva RT-PCR is an attractive alternative for the detection of SARS-CoV-2 in adults with much less known in children.

**Methods:** Children and adolescents with symptoms suggestive of COVID-19 were prospectively enrolled in a comparative clinical trial of saliva and nasopharyngeal (NP) RT-PCR between November and December 2020. Detection rates and sensitivities of saliva and NP RT-PCR were compared. Participants with discordant NP and saliva RT-PCR results including viral load (VL) were also analyzed.

**Result:** Out of 405 patients enrolled, 397 patients had two tests performed. Mean age was 12.7 years (range 1.2-17.9). Detection rates were 22.9% (95%CI 18.8-27.1%) by saliva RT-PCR, 25.4% (21.2-29.7%) by NP RT-PCR, and 26.7% (22.4-31.1%) by any test. The sensitivity of saliva was 85.2% (78.2-92.1%) when using NP as the gold standard; in contrast, when saliva was considered the gold standard, the sensitivity of NP was 94.5% (89.8-99.2%).For a NP RT-PCR VL threshold of ≥10^3^ and ≥10^4^ copies/ml, sensitivity of saliva increases to 88.7% and 95.2% respectively. Sensitivity of saliva and NP swabs was respectively 89.5% and 95.3% in patient with symptoms less than 4 days (p=0.249) and 70.0% and 95.0% in those with symptoms ≥ 4 to 7 days (p=0.096). The 15 patients who had an isolated positive NP RT-PCR were significantly younger (p=0.034), had a lower NP VL (median 5.6×10^3^ vs 3.9×10^7^, p<0.001), and were not able to drool saliva at the end of the sampling (p=0.002). VLs were significantly lower with saliva PCR than with NP RT-PCR (median 8.7 cp/ml x10^4^; IQR 1.2×10^4^-5.2×10^5^; vs median 4.0×10^7^cp/ml; IQR 8.6×10^5^-1.x10^8^; p<0.001).

**Conclusion:** Saliva PCR shows diagnostic performances close to NP RT-PCR for SARS-CoV2 detection in most symptomatic outpatient children and adolescents.

## Introduction

Diagnosis for SARS-CoV-2 is pivotal in the management of Coronavirus disease 2019 (COVID-19). Accurate and prompt testing of symptomatic children is the foundation for public health decision-making and implementation of appropriate measures including isolation and quarantine ^1^.

The Infectious Diseases Society of America (IDSA) guidelines recommend testing for SARS-CoV-2 by RT-PCR on specimens including NP swabs, mid-turbinate swabs, nasal swabs or saliva swabs, NP swabs being referenced as the gold standard specimen ^2,3^. However, NP swab collection is an unpleasant procedure that can sometimes result in mucous membrane erosions with nasal bleeding, especially in children ^4^. In addition, NP swabs require trained staff for collection and can be hampered by supply shortages in swabs and transport media. Saliva has already been reported as an attractive alternative for the detection of SARS-CoV-2 and other respiratory viruses such as Influenza in adult patients ^3,5^. A recent meta-analysis, reported a sensitivity of 83.2% (95%CI 74.7-91.4%) and 84.8% (95%CI 76.8-92.4%) for saliva and NP samples respectively ^6^.

The use of NP swabs for SARS-CoV-2 detection limits the widespread screening of children ^7^. Given the overlap of symptoms caused by SARS-CoV-2 and other respiratory viruses, children qualify for SARS-CoV-2 testing very often. As a result, a simple specimen collection such as saliva that avoids unnecessary discomfort is particularly attractive in children. Saliva collection has also a practical advantage since it can be performed quickly by the patient itself or by the caregiver. Altogether, the use of saliva specimens would speed up the collection process ^7^.

Pediatric evidence for the use of saliva specimens by RT-PCR for detection of SARS-CoV-2 remains weak with sensitivities varying from 52.9% up to 85.0% reported from small sample sizes when using NP as the gold standard. ^8,9^ This ancillary study of the adult RADICO project ^10^ aims to prospectively compare the paired saliva and NP samples collected from symptomatic children consulting in outpatient settings for the detection of SARS-CoV-2. The secondary objectives were to compare discordant NP and saliva RT-PCR findings as well as their VL.

## Methods

### Study design, setting and populations

This study is an observational prospective multicenter comparative study. Children aged 1 month to 18 years were recruited from two different outpatient clinics in Lausanne (Montétan screening site, Department Mother-Woman-Child; Lausanne University Hospital and Vidy-Med Pediatric Emergency Center) when presenting with symptoms compatible with COVID-19 according to national guidelines(check.bag-coronavirus.ch/screening)^11^. Children aged 12 and over who reported at least one of the following symptom including fever, respiratory symptoms such as cough, throat pain, dyspnea or thoracic pain, anosmia, dysgeusia or a least one minor symptom and close contact with a documented COVID-19 case were invited to be tested for SARS-CoV-2 (heck.bag-coronavirus.ch/screening). Testing criteria were more restrictive for children under 12 years of age^11^ (www.coronabambini.ch). For children without known contact with a SARS-CoV-2 positive case, testing criteria included a new onset of fever (>38.5 C), a severe cough associated with a bad general condition or the same symptoms persisting over 3 days in those with a good general condition. In addition, children with a good clinical condition presenting with a new onset of fever or a severe cough associated with other COVID-19 compatible symptoms such as gastrointestinal symptoms, myalgia, headaches, or anosmia/ageusia were also tested for SARS-CoV-2. Children with any symptom exposed to a confirmed SARS-CoV-2 positive index case were also tested for SARS-CoV-2.

Informed consent from the legal guardians or adolescents ≥14 years were mandatory for inclusion. Exclusion criteria included hospitalized children, those requiring anticoagulation, and children with a documented past SARS-CoV-2 infection.

This study was approved by the ethics committee of Canton de Vaud (CER-VD 2020-02269) and conducted in accordance with the principles of the Declaration of Helsinki, the standards of Good Clinical Practice, and Swiss regulatory requirements.

### Study procedures

Saliva specimens for RT-PCR analyses were collected either by a healthcare professional, the patient itself, or its caregiver under supervision, following a standard procedure that included swabbing of the oral mucosa and drooling saliva in a tube when possible ^10,12^. The healthcare professional collected concomitantly one NP swab for RT-PCR. Saliva and NP samples collected in viral transport media (VTM) were sent the same day or the next morning to the molecular diagnostics laboratory for RT-PCR analyses.

General information including age, gender, type and duration of symptoms, and information on the quality of the saliva sample were collected by the healthcare worker on an electronic case report form (REDCap®). The results of both NP swabs and saliva samples were next reported in REDCap® by the study investigators.

### SARS-CoV-2 RT-PCR, cycle thresholds (CT) and viral load (VL) quantification

SARS-CoV-2 RT-PCR were performed using an in-house RT-PCR on the automated molecular diagnostic platform targeting the E gene ^13–15^ or using the SARS-CoV-2 test of the Cobas® 6800 instrument (Roche, Basel, Switzerland)^16^. The cycle threshold (cycle when the RT-PCR was positive, i.e above the threshold of fluorescence) was provided automatically by the instruments by using the default parameters. Viral load was then obtained by converting (CT) of the RT-PCR instruments, using the formula logVL=-0.27Ct+13.04, as previously reported ^17,18^.

### Outcomes

The primary outcome was the proportion of SARS-CoV-2 positive children detected from saliva samples and NP swabs by RT-PCR assays. The secondary outcome was the viral loads of SARS-CoV-2 measured by RT-PCR assays on saliva and NP samples

### Statistical analysis

The estimated sample size was 50 positives among 500 cases tested to have a precision of +/-2% on the detection rate if the latter was 20%. All patients having a result available for the 2 tests were included in the study analysis population. Standard descriptive and comparative statistics were performed. The Chi-square test was used to compare categorical variables between groups, as appropriate for comparison between proportions. A Student t-test was used to compare continuous variables when the normality of the distribution was visually accepted. For skewed data, we derived medians and used the Mann–Whitney method for comparison. We estimated the detection rate and sensitivity of each test with 95% CIs. The sensitivity of saliva and NP samples was first calculated by using each other as the gold standard. Next, a composite gold standard combining any positive RT-PCR result reported from saliva and/or NP swabs was used to determine the sensitivity of both samples. Stratified subgroup analyses for different age groups were conducted. Participants were divided into 3 age groups: 0-6, ≥6-12, and ≥12 years of age and 3 symptom duration groups: 0-3, ≥4-7, and >7 days. Post-hoc analyses using the Bonferroni correction were performed between age groups (age group effect: p <0.017 (0.05/3)) that presented statistically significant different detection rates and sensitivities. Univariable linear regression was used to compare the clinical correlates (duration of symptoms and age of patients) to NP or saliva sample viral loads. Statistical analyses were computed using R software, v 3.6.1, and the 2019 R Studio interface (R Studio Team, Boston, MA).

## Results

### Patient characteristics

Eight hundred and seventy-eight children and adolescents were screened between November 4^th^ and December 12^th^, 2020 for SARS-CoV-2. Among them, 405 children were included in this study, of whom the 397 who had both NP and saliva samples collected were included in the analyses. The mean age was 12.7 years (Standard Deviation (SD) 3.8, range 1.2 to 17.9). One hundred and ninety-two patients (48.3%) were females. The characteristics of the patient population stratified by SARS-CoV-2 positive and negative results are summarized in Table 1.

**Table 1.**
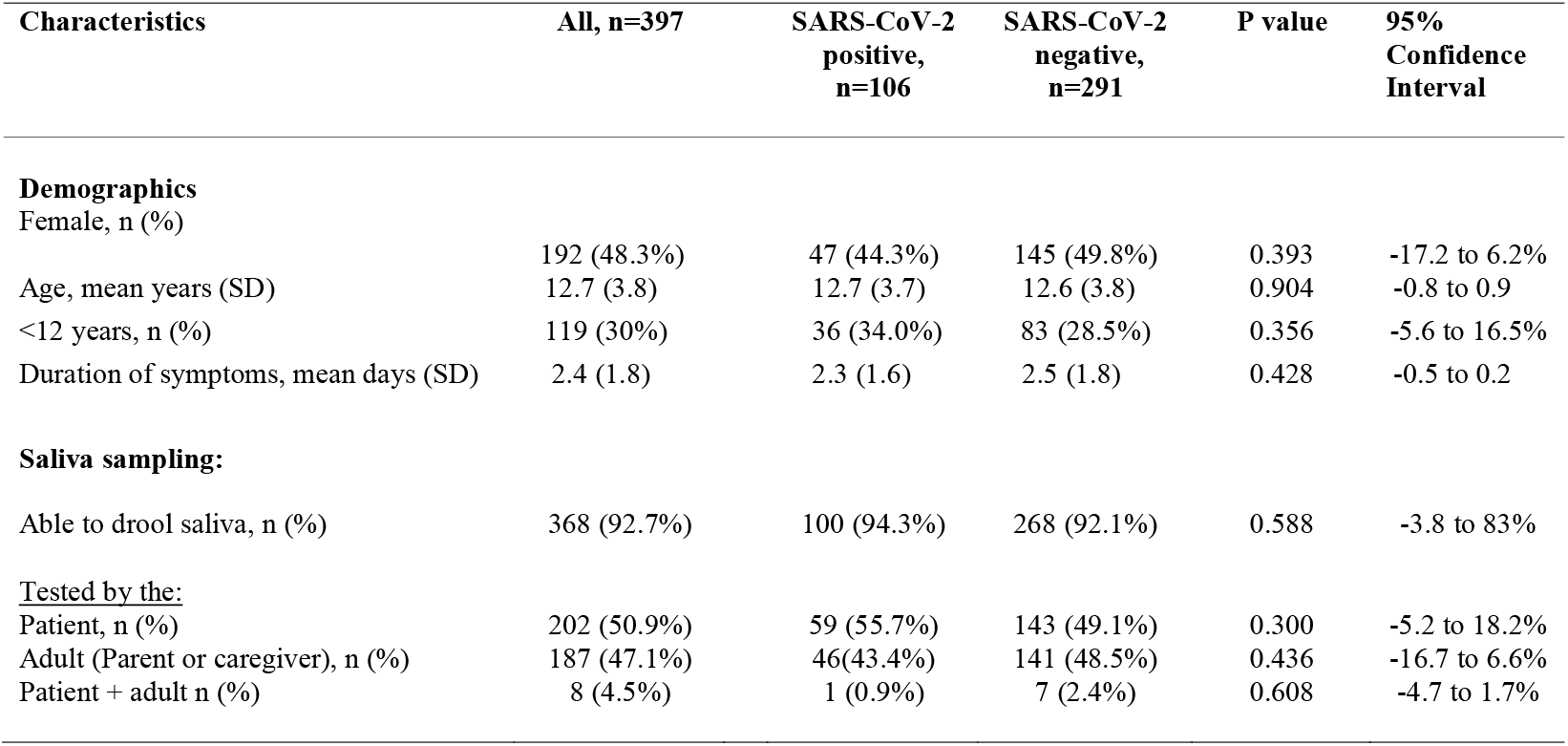
Characteristics stratified by SARS-CoV-2 positive or negative results Legend: NP: Nasopharyngeal swab, SD= Standard Deviation

The mean time between the onset of symptoms and the collection of the NP and saliva samples was 2.4 days (SD 1.8, range 0 to 10) with no significant differences reported in the duration of symptoms between children under 12 years of age and those aged 12 and over (p=0.697). A vast majority of children of ≥12 years of age presented at least one major symptom (89.9%), mostly sore throat (68.6%), cough (49.5%), fever (25.6%), dysgeusia (13.4%) and anosmia (10.5%). The remaining 10.1% (28/278) suffered from at least one minor symptom while having close contact with a documented COVID-19 case. From children <12 years of age, 81.5 % (97/117) presented with fever (47.9%) and/or a severe cough (52.1%) associated at least with a bad general condition, other manifestations suggestive of COVID-19 or symptoms lasting more than 3 days. The remaining 18.5% (22/117) suffered from at least one symptom suggestive of COVID-19 and had close contact with a documented COVID-19 case.

### Detection rates of NP RT-PCR and saliva RT-PCR

Of the 397 participants included in the analyses, 91 (22.9%; 18.8-27.1%) tested positive by saliva samples, 101 (25.4%; 21.2-29.7%) by NP swabs, and 106 (26.7%; 22.4-31.1%) by any of the 2 samples. Detection rates were equivalent for both NP and saliva specimens (−8.7 to 3.7%, p=0.457). Respectively 15 and 5 children were detected positive only on NP swabs or saliva specimens. The detection rates by age categories (0-6, ≥6-12, and ≥12 years of age) were significantly different for saliva samples (p=0.007) but not for NP swabs (p=0.070). Yet, this remained only statistically different in post-hoc analyses between the age groups of 0-6 years and ≥6-12 years with a higher detection rate among children of ≥6-12 years of age (3.2% vs 30.7 %, p=0.004) (Figure 1)

**Figure 1:**
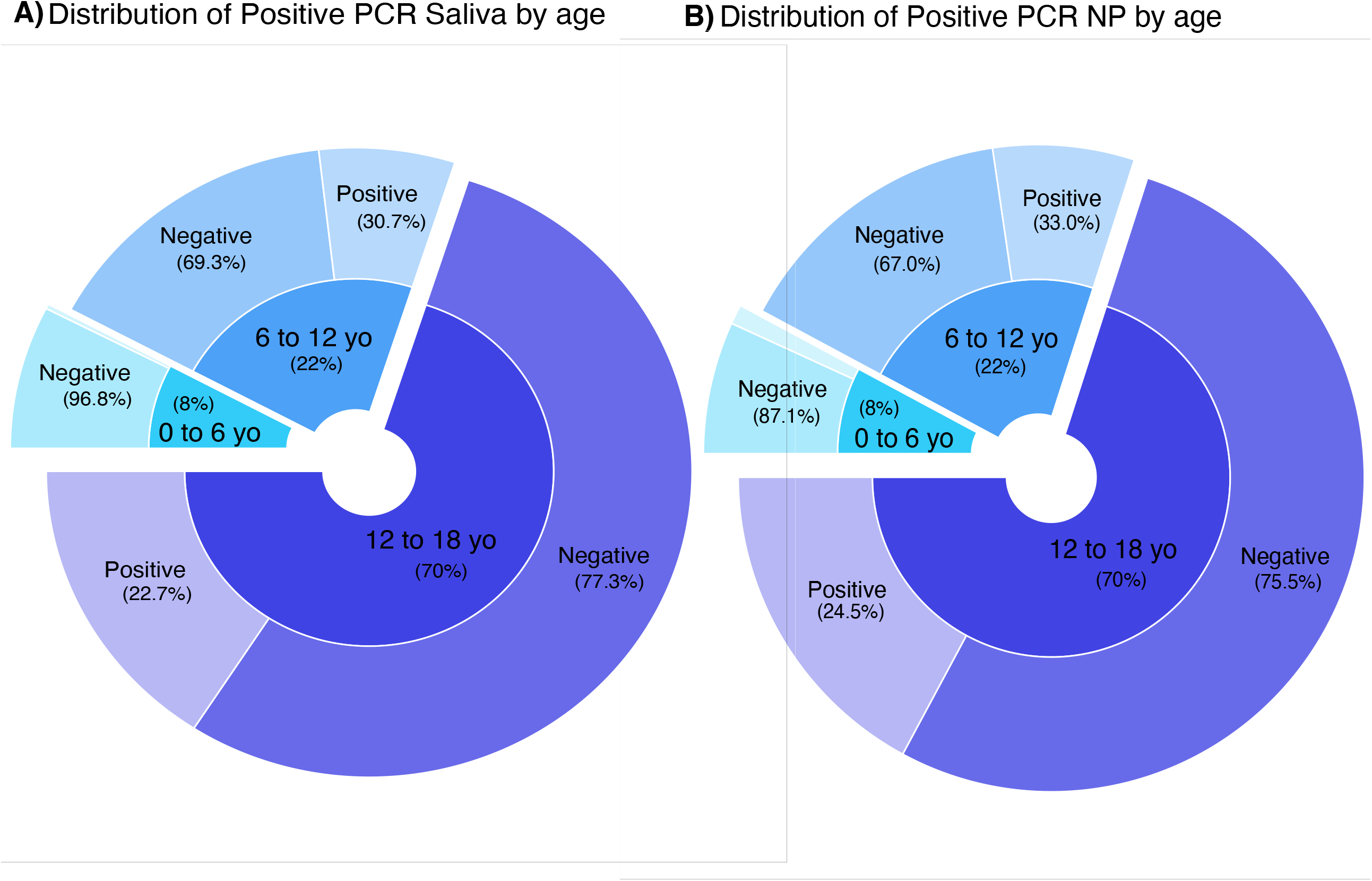
Detection rate for NP and saliva PCR by age categories. NP= Nasopharyngeal

### Diagnostic test performance (sensitivity, specificity) of NP RT-PCR and saliva RT-PCR

Using NP as the gold standard, the sensitivity of saliva was 85.2% (95%CI 78.2-92.1%); in contrast, when saliva was considered the gold standard, the sensitivity of NP was 94.5% (95%CI 89.8-99.2%) (p=0.058). The sensitivity of saliva RT-PCR was dependent on NP VLs and was maximal with a viral load of 10^6^ cp/ml. (Table 2 and Figure 2). When using the composite reference as the gold standard, the respective sensitivity of saliva and NP swabs was 85.9% (95%CI 79.2-92.5%) and 95.3% (95%CI 91.3-99.3%) (p=0.034). When stratified by age groups, the respective sensitivity of saliva and NP swabs was 89.9% and 97.1% in children aged ≥12 years and 84.4% and 90.6% in children ≥ 6 to 12 years of age. In children under 6 years of age, 4 patients were detected positive from NP swabs, whereas only one child was documented positive from saliva. As a result, the reported sensitivity was only 25% for saliva PCR and 100% for NP swabs in this subgroup. The reported sensitivity was significantly different between age groups for saliva samples (p=0.0001) but not for NP swabs (p=0.320). Yet, in post-hoc analyses, sensitivity for saliva remained statistically different only between the 0-6 year subgroup compared to the one including children of 12 years of age and onwards (25% vs 89,9%, p=0.003). When stratified by the duration of symptoms, the respective sensitivity of saliva and NP swabs was 89.5% and 95.3% in patient with symptom duration < 4 days (95%CI - 14.8 to 3.2%, p=0.249) and 70.0% and 95.0% in those with symptom duration (95%CI -52.2 to 2.2%, p=0.096) ≥ 4 to 7 days. Only 3 patients had symptoms above 7 days and all were tested negative from both samples.

**Table 2.** Sensitivity of saliva RT-PCR compared to NP RT-PCR stratified by viral load thresholds. Legend: NP: Nasopharyngeal swab, VL= viral load

|  |  | **Sensitivity** |  |  |  |
| --- | --- | --- | --- | --- | --- |
| **VL ≥** | **VL <1e+3** | **VL ≥1e+3** | **VL ≥1e+4** | **VL ≥1e+5** | **VL ≥1e+6** |
| **Saliva PCR Sensitivity** | 85.1% | 88.7% | 95.2% | 97.3% | 98.4% |

**Figure 2:**
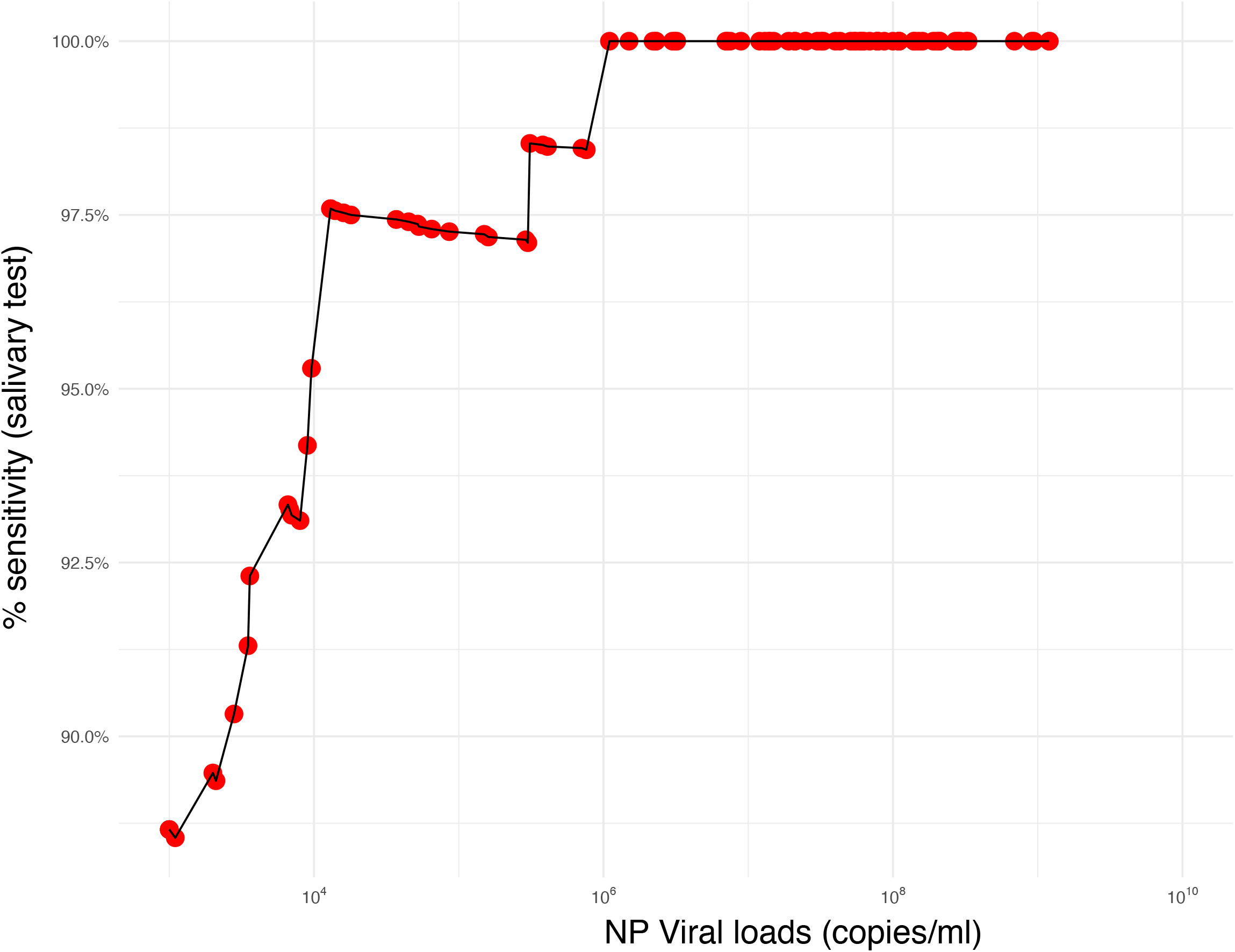
Sensitivity of saliva PCR in relation to NP viral loads. NP= Nasopharyngeal

### Viral loads

Figures 3A and 3B display the distribution of VLs and CT by analyzed specimen. VLs documented from saliva were significantly lower compared to those reported from paired NP swabs (median 8.7 cp/ml x10^4^; IQR 1.2×10^4^-5.2×10^5^; vs median 4.0×10^7^cp/ml; IQR 8.6×10^5^-1.x10^8^; p<0.001, 95CI: -4.5×10^2^ to - 7.7×101). VLs measured from saliva increased significantly with age but not with the duration of symptoms (Figure 4A/B). VLs measured from NP were not affected by age. (Figure 4A).

**Figure 3:**
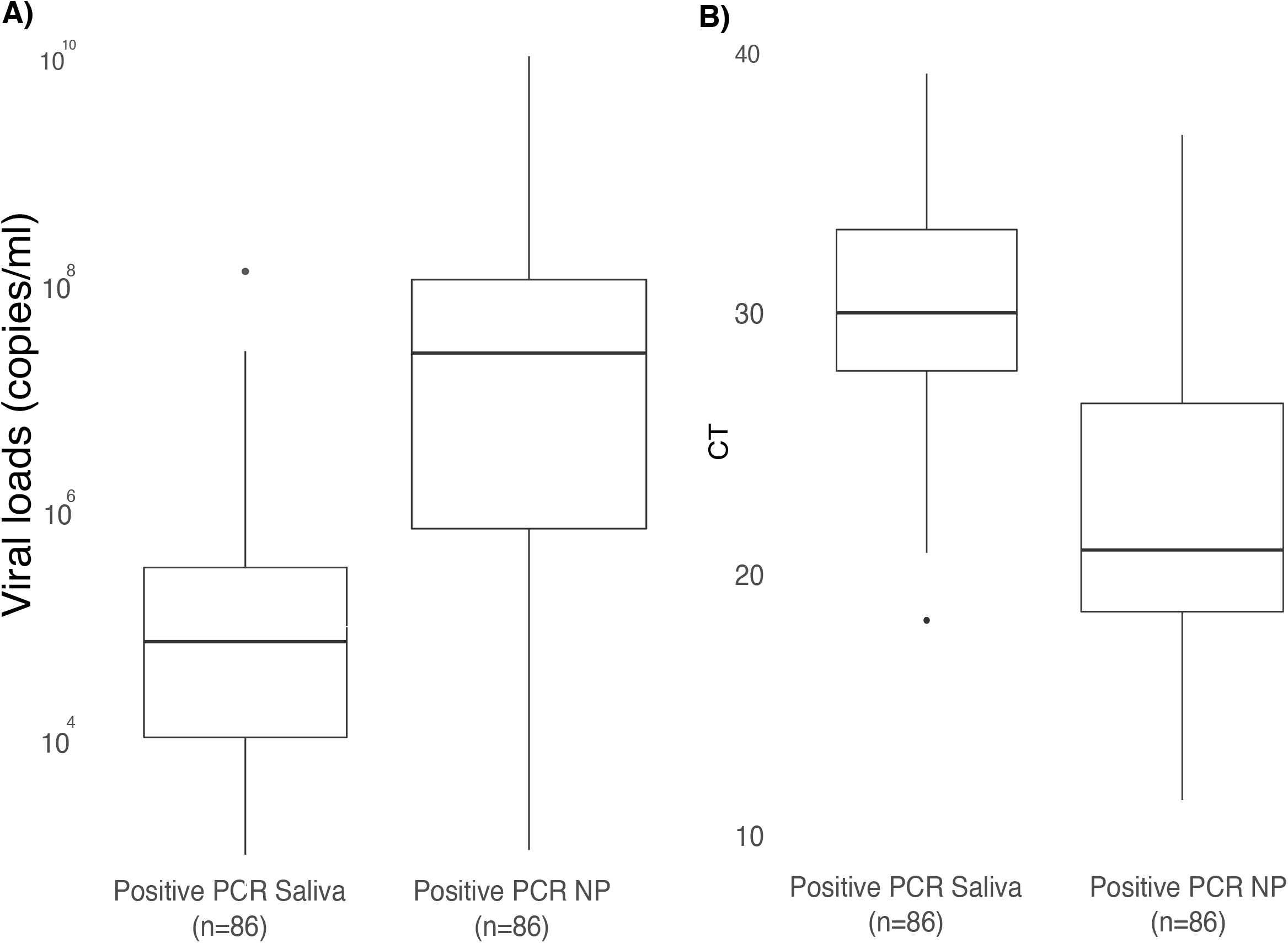
Distribution of viral load and CT according to the type of sample. NP= Nasopharyngeal

**Figure 4:**
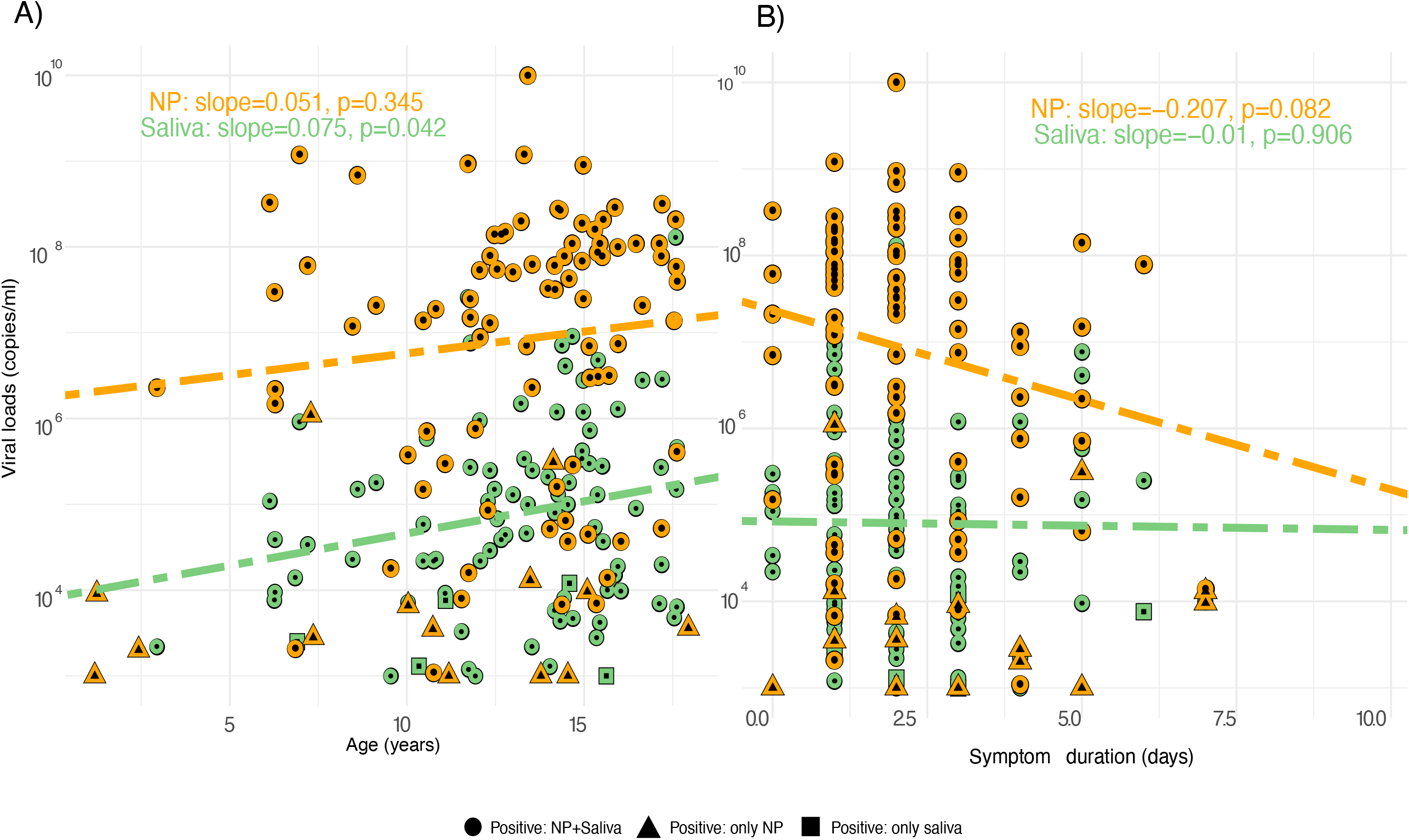
Relation of viral loads to symptom duration and age.

### Comparison of discordant NP and saliva RT-PCR results

Table 3 displays the characteristics of the 15 children with NP swabs only positive compared to the 86 documented positive from both NP swabs and saliva samples. Children with isolated SARS-CoV-2 positive NP swabs were significantly younger (p=0.034) and with significantly lower NP VLs (median 5.6×10^3^ vs 3.9×10^7^, p<0.001) compared to those with both saliva and NP positive specimens. In addition, they were not able to drool at the end of saliva collection (p=0.002). Variables such as the duration of symptoms or the person who performed the procedure did not affect the above findings. The 5 patients who had isolated positive saliva samples were all males, had a median age of 11.1 y (10.3-14.6), and presented with median VLs of 4.0×10^3^ cp/ml (1.1×10^3^-8.8×10^3^).

**Table 3.** Characteristics stratified by the concordance of positive NP and saliva RT-PCR findings Legend: NP: Nasopharyngeal swabs, VL= viral loads, IQR= interquartile range IQR= interquartile range

| Characteristics | Negative saliva PCR,<br>NP PCR + n=15 | NP and<br>saliva<br>PCR +,<br>n=86 | P value | 95%<br>Confidence<br>interval |
| --- | --- | --- | --- | --- |
| Age, median (IQR) | 11.2 (7.3-14.0) | 14.1 (11.8-15.4) | **0.034** | -4.7 to -0.2 |
| Female, n (%) | 9 (60.0%) | 45 (52.3%) | 0.788 | -23.2 to 38.5% |
| Duration of symptoms, median,<br>(IQR) | 3 (1.5-4.5) | 2 (1-3) | 0.094 | 0 to 2 |
| NP viral load, median (IQR) | 5.6x10<sup>3</sup><br>(1.5x10<sup>3</sup>-1.0x10<sup>4</sup>) | 3.9x10<sup>7</sup><br>(8.6x10<sup>5</sup>-1.0x10<sup>8</sup>) | **<0.001** | -1.5x10<sup>4</sup>to -4.6x10<sup>2</sup> |
| **<u>Tested by the:</u>** |  |  |  |  |
| Participant, n (%) | 7 (46.7) | 50(58.1%) | 0.586 | -42.7 to 19.8% |
| Adult (Parent or caregiver), n (%) | 8 (53.3) | 35 (40.7 %) | 0.529 | -18.6 to 43.8% |
| Patient + Adult, n (%) | 0 (0.0 %) | 1(1.1%) | 1.000 | -4.6 to 2.3% |
| Able to drool saliva, n (%) | 11(73.3%) | 84(97.7%) | **0.002** | -50.9 to 2.2% |

IQR= interquartile range

## Discussion

The overall detection rate for SARS-CoV-2 between saliva and NP specimens by RT-PCR was comparable. However, a significantly lower sensitivity was reported from saliva specimens compared to NP swabs when compared to any positive test. Yet, the sensitivity of saliva increased with VLs and was equivalent to NP swabs when using a VL threshold of ≥10^4^ copies/ml. The procedure of saliva collection and younger age affected the detection of SARS-CoV-2 in saliva. In contrast, the duration of symptoms over 7 days of onset did not significantly affect the sensitivity of SARS-CoV-2 detection in saliva.

With more than twice as many children included ^8^, this study is by far, the largest cohort reporting on the performance of saliva specimens in children. As supported by another study including children ^8^ and other studies conducted in adults ^10,19^, a detection rates slightly lower of SARS-COV-2 were reported from saliva as compared to NP specimens, yet still supporting the use of saliva as an alternative to NP swabs in children. Moreover, our findings are in line with the RADICO study conducted in symptomatic adults, that used the same saliva collection approach ^10^. In contrast, other studies indicated a poor concordance between NP and saliva specimens ^9,20,21^, likely as a result of using different sampling procedures that did not necessarily include saliva drooling. In addition, the inclusion of adults and children with a variable duration of symptoms upon testing and asymptomatic patients possibly limited their conclusions.

The sensitivity of RT-PCR assays reported from saliva was lower compared to NP swabs and the gold standard test, albeit only reaching statistical difference when compared to any positive test. This difference in sensitivity is likely explained by a 2 log lower VL detection in saliva compared to NP samples as evidenced elsewhere ^9,10,22^. Furthermore, SARS-CoV-2 detection in saliva is dependent on viral loads and reaches an equivalent sensitivity to NP swabs for NP VL thresholds of 10^4^ copies/ml. Recent studies ^23–25^ suggest that no cultivable viable virus is detected from patients with VLs under the threshold of 10^4^ copies/ml. Altogether ours and others findings suggest ^8^ that children detected SARS-CoV-2 negative from saliva samples presented VLs under the threshold of 10^4^ copies/ml from their NP and were thereby potentially less contagious. The use of saliva as an alternative to NP for SARS-CoV-2 detection in children would thus limit quarantine measures to most contagious children ^26^.

Younger age significantly affected the detection rates and the sensitivity of SARS-CoV-2 in saliva as supported by a significantly reduced sensitivity of saliva documented among children under 6 years of age compared to those of 12 years of age and onwards. Furthermore, children who only detected positive from their NP swabs were significantly younger compared to those documented SARS-CoV-2 positive from both specimens. The significant linear association documented between age and VLs supported that lower VLs documented among younger children likely affected the sensitivity of saliva for SARS-CoV-2 detection. Yet, children under 6 years of age only represented 8% of our cohort, thereby limiting our conclusions. In addition, the incomplete procedure for saliva collection observed among younger children might have impacted the sensitivity, as most of the 15 children only documented SARS-CoV-2 positive from their NP swabs were not able to drool. Yet, data from the RADICO study reported no impact of drooling on the sensitivity of saliva nor the VL count ^10^. Whether drooling affects the sensitivity of saliva remains an open debate and should be investigated in future pediatric studies.

Furthermore, drooling into the tube also raises biosafety concerns. The edge of the tube can be contaminated thereby questionning the use of precautionary measures. Duration of symptoms over 7 days of onset did not significantly affect the sensitivity of SARS-CoV-2 detection in saliva as already supported in other adult ^19^ and paediatrics ^9^ studies.

Strengths of the current study include the largest pediatric sample size collected so far in addition to the detailed prospectively collected information. Limitations are predominantly related to the inclusion of outpatients and not hospitalized nor asymptomatic children, which might affect the generalizability of our findings. Furthermore, young children were under-represented and only a few of them were detected SARS-CoV2 positive, thus limiting extrapolation to this age-group. In addition, our study was conducted during a high prevalence of SARS-CoV-2 (up to 30%), thereby affecting our positive predicted values. Finally, our study was conducted before the introduction of 501Y mutants in Switzerland, which currently represent 8 to 15% of the analyzed samples. As such, we were not able to compare the performance of mutant typing in saliva vs NP. Yet, recent evidence supports that the lower VLs detected from saliva limit mutant typing analyses ^27^.

## Conclusion

In conclusion, saliva is a reliable alternative for SARS-CoV-2 detection among symptomatic children. Saliva collection being a non-invasive easy procedure will facilitate large-scale screening in children and thus provide more evidence on the impact of SARS-CoV-2 in children.

## Data Availability

All data referred in this manuscript ara available

## Funding

The saliva PCR were paid for by the cantonal health authorities.

## Acknowledgments

We thank Marion de Vallière and Maria Daniela Garrido for help in patient recruitment, the patients for participation in this study, and the health authorities of the Canton de Vaud for their support. We would like to thank the Vidy-Med team and the Montétan screening site teams for the recruitment of participants. We thank all the team of the Laboratory of Molecular Diagnostics of the Institute of Microbiology and Anne Tabard-Fougère for her help in the statistical analyses and figure elaboration.

## Conflict of interest

None

## Notes

### Competing Interest Statement

The authors have declared no competing interest.

### Clinical Trial

NCT04613310

### Author Declarations

CER-VD 2020-02269

